# Pulmonary exacerbations in patients with genetically confirmed PCD: A prospective observational multicentre study

**DOI:** 10.1101/2025.11.18.25340347

**Authors:** Pinelopi Anagnostopoulou, Panayiotis Kouis, Dilber Ademhan Tural, Sarah Altaraihi, Simona Basilicata, Melissa Borrelli, Athina Christofidou, Hatice Nur Coemert, Renato Cutrera, Nagehan Emiralioglou, Ela Erdem, Yasemin Gokdemir, Serena Gracci, Elpis Hatziagorou, Simone Helms, Bülent Karadag, Debora Maj, Marcus A. Mall, June K. Marthin, Nicos Middleton, Antonio Moreno-Galdó, Ugur Özçelik, Massimo Pifferi, Johanna Raidt, Jobst Röhmel, Sandra Rovira-Amigo, Francesca Santamaria, Ioannis Tsiolakis, Nicola Ullmann, Niklas Ziegahn, Kim G. Nielsen, Heymut Omran, Panayiotis Yiallouros

## Abstract

**Background:** Primary ciliary dyskinesia (PCD) is a rare genetic disorder characterized by deficient mucociliary clearance and development of chronic lung disease. Pulmonary exacerbations (PEx) in chronic lung diseases increase morbidity and lung function decline, but their frequency in PCD has been understudied. We aimed to prospectively determine the annual frequency of PEx in PCD and identify related risk factors.

**Methods:** In a multicentre, observational study conducted in 11 centres from seven countries, we prospectively collected data from a well-described patient cohort with genetically confirmed PCD over a year via monthly telephone questionnaires and clinical visits. We assessed the annual PEx frequency using three definitions: i) clinical definition 1 (*Def-1*) when three out of seven clinical items were positive; clinical definition 2 (*Def-2*) if the patient started or changed antibiotic treatment; *self-reported* PEx by the patient). For paired statistical comparisons we used the Friedman test and the Wilcoxon matched-pairs test and tested their agreement (Cohen’s kappa statistics). We also determined related risk factors using a mixed-effects model.

**Results:** We recruited 271 individuals with PCD of all ages. Among patients with complete annual records (n=248), approximately 80% experienced at least one PEx per year, as assessed by the three definitions used. Self-reported PEx per year (median 2, interquartile range (IQR) 1-5) were higher (p<0.0001) than the PEx assessed by Def-1 (median 2, IQR 0-4) and Def-2 (median 1, IQR 0.25-3). Self-reported PEx had a substantial agreement with Def-1 [kappa (SE) =0.61 (0.05)] and a moderate agreement with Def-2 [kappa (SE) =0.51 (0.05)]. Female sex and autumn season were associated with significantly higher number of PEx, independent of the definition used. Increasing age was correlated with higher annual PEx frequency by Def-1.

**Conclusion:** In this multicentre study, we prospectively assessed the annual PEx frequency in patients genetically diagnosed with PCD, demonstrating the importance of the definition used in capturing the exacerbation burden of PCD, as well as the influence of sex, age and season on exacerbation frequency.

## INTRODUCTION

Primary ciliary dyskinesia (PCD) is a heterogeneous rare genetic disorder characterized by dysfunction of the motile cilia [1] [2]. Its prevalence is estimated to be 1:7500 people based on genome data, with high heterogeneity across countries [3] [4]. PCD mainly affects the respiratory system, causing defects in mucociliary clearance [5] [6] [7]. As the ciliated epithelium covers most of the respiratory tract, patients usually present with symptoms from the upper airways, such as chronic rhinosinusitis and conducting hearing impairment, and the lower airways, such as chronic productive cough and often bronchiectasis [2] [8], in consistency with the concept of united airways’ disease [9]. In addition, patients with PCD frequently suffer from pulmonary exacerbations (PEx), experiencing worsening of symptoms from the lower respiratory tract. These reflect an immune host response possibly due to the presence of microorganisms, which may require antibiotic treatment [10], and/or the impact of other noxious agents [11] [12]. PEx have been linked to increased morbidity and lung function decline [13] [14] [15].

The frequency of PEx in chronic lung diseases is an important determinant of the disease burden and is often used as a primary outcome in clinical trials [16] [17]. Data on PEx frequency in PCD are scarce [18] [19]. Additionally, the definition of PEx in PCD remains controversial. A few clinical trials have used the ‘‘physician’s or patient’s decision to initiate or change antibiotic treatment for respiratory symptoms” to define a PEx [20] [21] [13]. Recently, a consensus statement on PEx definition in patients with PCD, independent of age, has been published. It was developed by a multidisciplinary group of experts and patients using a Delphi process and was based on clinical characteristics [14]. To date no study has validated either definition of PEx using prospective real world patient data.

In this prospective, multicentre study, we performed monthly structured interviews and retrieved parallel clinical data from patients with PCD of all ages for at least 12 months. The primary aim was to determine the frequency of PEx within one year using the above definitions, as well as the patients’ personal perception of having an exacerbation. We also aimed to i) compare the annual PEx frequency using the different definitions, ii) identify determinants that increase the risk for PEx, and iii) study the seasonal distribution of PEx.

## METHODS

### Study design

This is a prospective, multicentre, observational study conducted in 11 centres within the ERN-LUNG PCD core from seven countries [22] (see Supplementary data).

### Study participants

We included patients of all ages with a confirmed diagnosis of PCD based on genetics [23], who participate in the International PCD Registry (NCT02419365) [24]. The study was approved by the ethics committee of the Westphalian Wilhelms–University of Muenster (Muenster, Germany; AZ 2011–270-f-S) and the local ethics committees at each centre. Patients and/or legal guardians signed an informed consent prior entry to the study.

### Genetic diagnosis

Participants’ genetic data were retrieved from the ERN-LUNG International PCD Registry. Genetic variants were evaluated according to published guidelines [25] [26] (see Supplementary data).

### Data Collection

#### Monthly structured telephone interviews using a standardised questionnaire

A trained healthcare professional conducted monthly a telephone interview with the patient or the caregiver for children ≤12 years, and completed the questionnaire developed for the needs of the study (Table 1) (further details in Supplementary data).

During the interview, the patient/caregiver was asked to provide specific dates for the items referred in Table 1. This information was later used to exclude overlap of exacerbations within two consecutive months. In this case, we counted the PEx in the first month that it appeared.

**Table 1:** Definitions of pulmonary exacerbation (PEx) based on questions included in the monthly questionnaire of the study, and respective responses; * the first question in the questionnaire; *^$^* the antibiotic treatment was either decided by a physician or self-administered by the patient/caregiver. For each positive answer to questions corresponding to Def-1 and Def-2 the patient had to give specific dates of duration

| Definition | Question (s) | PEx - determining response |
| --- | --- | --- |
| **Self-reported** | *'In the last month how many exacerbations of your disease have you had? By this I mean infections, bad attacks of your chest / worsening of your symptoms'\** | One or more exacerbations |
|  | i) *'During the last month did you experience increased cough?'* |  |
|  | ii) *'During the last month did you experience increased sputum?'* |  |
| **Definition 1**<br>**(Def-1)**<br>[14] | iii) *'During the last month did you experience dyspnoea?'* | Positive response in at least three questions |
|  | iv) *'During the last month did you start/change antibiotic treatment due to pulmonary symptoms?'* |  |
|  | v) *'During the last month did you experience malaise?'* |  |
|  | vi) *'During the last month did you experience hemoptysis?'* |  |
|  | vii) *'During the last month did you experience fever (body temperature > 38°C)?'* |  |
| **Definition 2**<br>**(Def-2)**<br>[15] [20] [21] | *'During the last month did you start/change antibiotic treatment due to pulmonary symptoms?'*<sup>\$</sup> | Positive response |

#### Clinical data

We used data (anthropometrics, sputum/pharyngeal swab microbiology, spirometry) collected in parallel routine and emergency clinical visits in each centre during the study period (further details in Supplementary data).

### Definitions of pulmonary exacerbation

We evaluated three definitions of PEx based on responses to specific questions or combination of questions included in the questionnaire as shown in Table 1.

<u>Self-reported PEx</u>: A positive (≥1) answer to the question ‘*In the last month how many exacerbations of your disease have you had? By this I mean infections, bad attacks of your chest / worsening of your symptoms’*. This was the first question in the questionnaire. Because a low number of questionnaires (3.6%) had an answer >1, for the analysis, we considered each response ≥1 equal to 1, which could have introduced a small degree of bias.

The above definition intends to cover the need to include patients’ personal perception of experiencing a PEx, which could be different from clinical definitions. Interestingly, in COPD, the frequency of exacerbations was found to be higher in studies that record the patients’ perception on PEx compared to those using clinical definitions for PEx [27] [28].

<u>Definition 1 (Def-1)</u>: Definition of pulmonary exacerbation if three or more of the following seven questions had a positive answer. If the answer was positive, we asked for specific dates that each item occurred.

i. *‘During the last month did you experience increased cough?’*
ii. *‘During the last month did you experience increased sputum?’*
iii. *‘During the last month did you experience dyspnoea?’*
iv. *‘During the last month did you start/change antibiotic treatment due to pulmonary symptoms?’*
v. *‘During the last month did you experience malaise?’*
vi. *‘During the last month did you experience hemoptysis?’*
vii. *‘During the last month did you experience fever (>38°C)?’* [14]

<u>Definition 2 (Def-2)</u>: A positive answer to the question ‘*During the last month did you start/change antibiotic treatment due to pulmonary symptoms?’*. The antibiotic treatment was either prescribed by a physician or self-determined by the patient/caregiver.

A separate question of the questionnaire asked if the patient required unscheduled admission to hospitals for the above problems, with recording of specific dates in case of a positive answer.

### Statistics

Statistical analysis was performed with Stata (Stata/SE 15.1, StataCorp LLC, TX, USA) and graphical presentation of the data with GraphPadPrism (Prism 9, V9.3.1, GraphPad Software LLC). To calculate the annual PEx frequency per person, we analysed the last 12 months of each patient’s participation in the study and included patients with at least 10 completed questionnaires during 12 consecutive months. Median (interquartile range, IQR) values are reported due to the non-normal distribution of the data. The PEx incidence rate was calculated as the total number of exacerbations recorded in the cohort/total person-years of observation (total number of questionnaires*30/365.25). We performed subgroup analysis pooling together genes predicted to cause abnormal (Class I) or normal (Class II) ciliary ultrastructure, as previously described [29] [30]. Other subgroup analyses included children vs adults, age-defined pediatric groups (children 0-12 years where a caregiver provided monthly data, and children 12.1 to 17.9 years that self-provided monthly data), as well as geographical groups (Northern/Central Europe vs Southern Europe). For the assessment of seasonal/monthly distribution, original monthly data from all participants were used.

Paired comparisons between the three PEx definitions were performed using the Friedman test and the Wilcoxon matched-pairs test. The Mann-Whitney test was used for unpaired comparisons between different patient subgroups and p-values were adjusted for multiple testing using the Benjamini-Hochberg Procedure (FDR correction). The Cohen’s kappa statistics was used to assess the agreement between definitions [31]. A mixed-effects model with a random effect for the centre was used to evaluate the association of the exacerbation frequency with patients’ characteristics, i.e. sex, age, FEV_1_ z-score and microbiology results. To identify the most relevant parameters to include in the mixed effects model, separate univariable linear regressions were performed for each parameter and variables with p<0.1 were included in the final model. Seasonal comparison was performed using odds ratios, considering that all centres are located in the northern hemisphere. A p-value <0.05 was assumed to indicate statistical significance.

## RESULTS

### Study participants

In total, we performed 3579 monthly structured interviews (median 12, range 1-25) in 271 people with PCD (56.8% males), median age 17 years (range 0-72). Participants’ age distribution is shown in Table 2. The median of the median FEV_1_ z-scores during the study was -1.14 (range -4.7; 2.5), with 66.5% of the participants having FEV_1_ within normal range (≥5^th^ percentile z-score) (Table 2) [32]. Among participants with available microbiology data (n=210), the most commonly isolated microorganisms were *Pseudomonas aeruginosa*, *Hemophilus influenzae* and *Methicillin-sensitive Staphylococcus aureus* (Table 2).

**Table 2:**
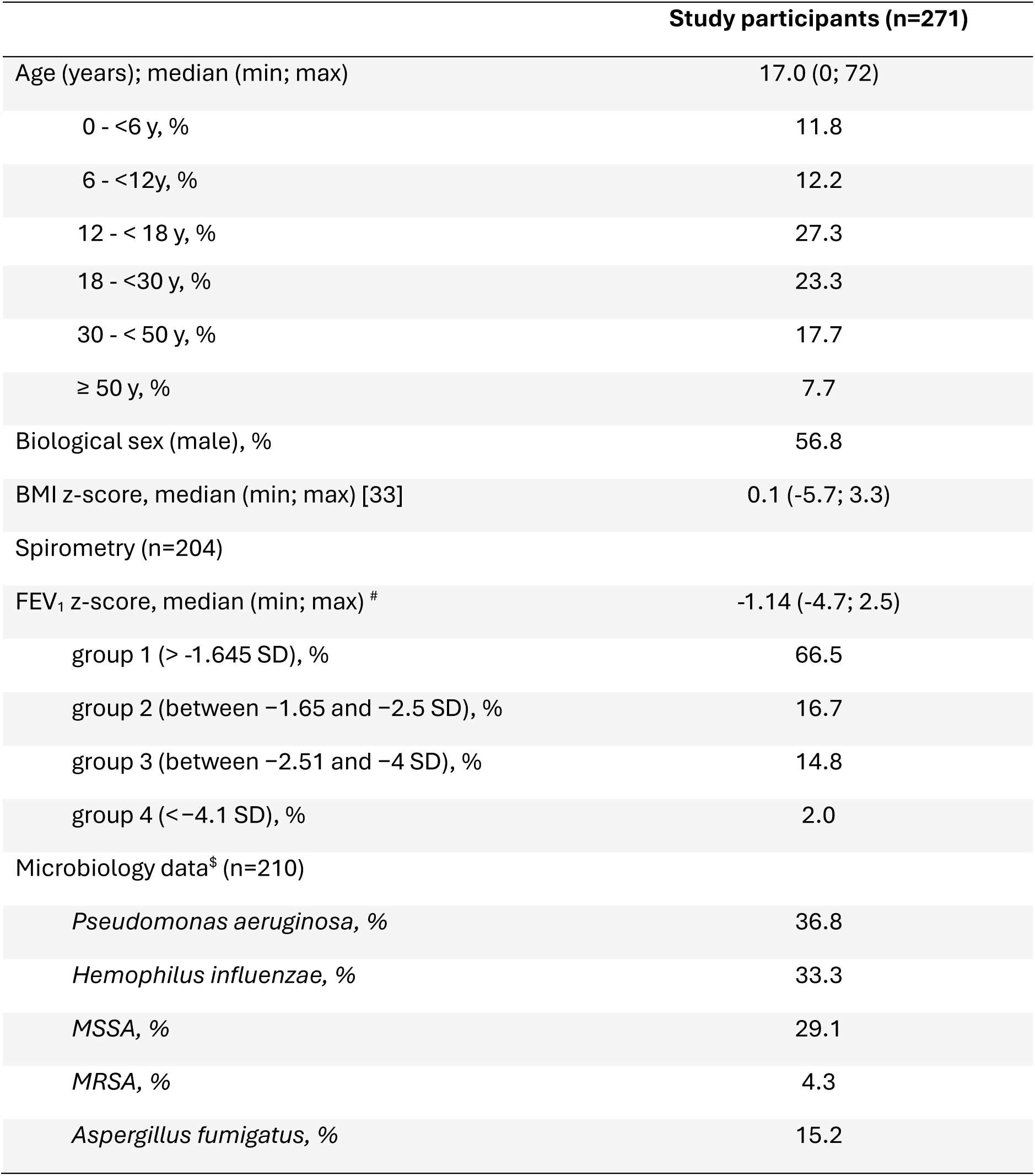
Demographics, age distribution, spirometry and microbiology data of the study participants; BMI: body mass index, FEV_1_: forced expired volume at 1 sec, SD: standard deviation, *MSSA; Methicillin-sensitive Staphylococcus aureus, MRSA: Methicillin-resistant Staphylococcus aureus*. ^#^ Median FEV_1_ during the study period; ^$^ At least once positive culture during the study period.

#### PCD diagnostics

Among 36 PCD-related genes identified in this population, the most frequent were *DNAH5* (n=61), *CCDC40* (n=34), *DNAH11* (n=32), *CCDC39* (n=18) and RSPH9 (n=11) (Supplementary Figure S1).

### Annual frequency of pulmonary exacerbations

Data from 248 patients with at least 10 completed questionnaires during 12 consecutive months were used to calculate the annual frequency of PEx per person. The annual frequency of self-reported PEx per person (median 2, IQR 1-5) was significantly higher than that recorded with Def-1 (median 2, IQR 0-4; p<0.0001) and with Def-2 (median 1, IQR 0.25-3; p<0.0001). There was also a significant difference in PEx per person per year assessed with Def-1 and Def-2 (p=0.019) (Fig. 1a). The incidence rate per year was 3.32 for the self-reported exacerbations, 2.51 for Def-1 and 2.06 for Def-2. In total, 207 (83.5%) participants experienced at least one PEx per year as defined by clinical definitions (Def-1 and/or Def-2), while the same percentage of participants, but not necessarily the same individuals, experienced at least one self-reported PEx per year (Figure 1b).

**Figure 1:**
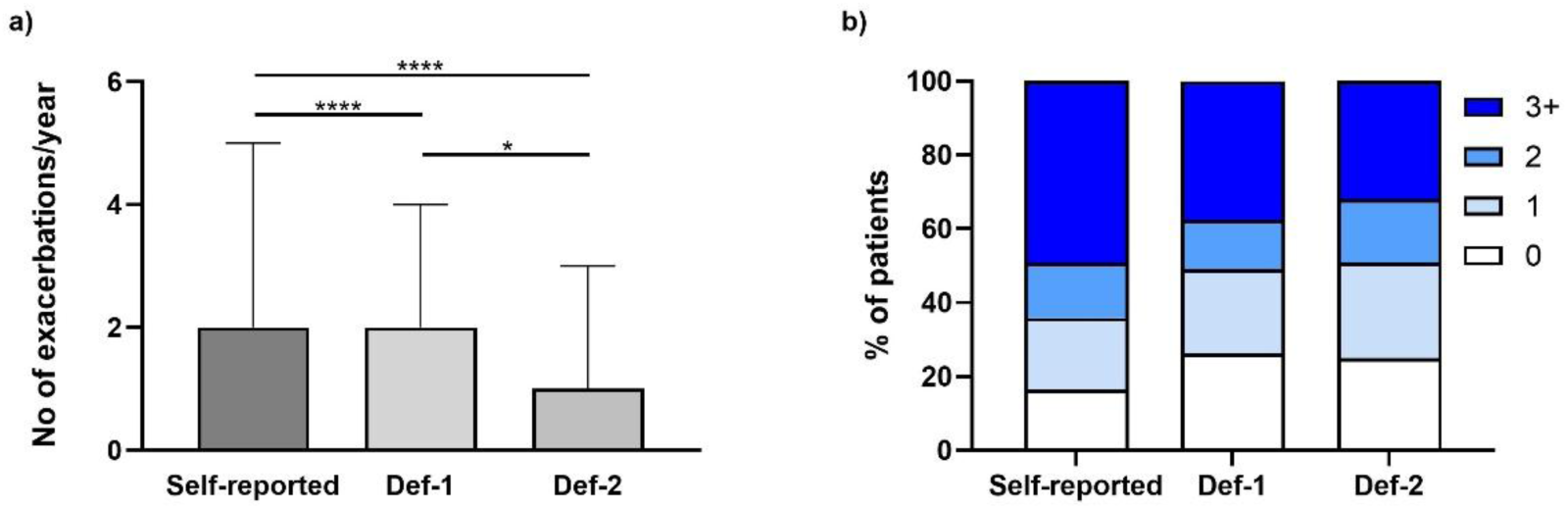
a) Annual frequency of pulmonary exacerbations, as recorded by self-reports (Self-reported), Definition 1 (Def-1) and Definition 2 (Def-2) in n=248 patients with genetic confirmation of PCD and annual records. Number (No) of exacerbations/year are shown as median (interquartile range). Friedman test and Wilcoxon-matched pairs test were used for statistical analysis; **** p<0.0001, *p=0.01. b) Percentage of patients reporting 0, 1, 2 and 3 or more (3+) exacerbations per year, as recorded by self-reports (Self-reported), Definition 1 (Def-1) and Definition 2 (Def-2)

### Hospitalisation rate for PEx

In total, 78 questionnaires reported hospitalisation for a PEx in the past month. Considering that hospitalisation included antibiotic IV treatment, this number corresponds to 12.9% of all reported exacerbations by Def-2 (N=604 exacerbations).

### Agreement between different definitions of PEx

We found a moderate agreement between Def-1 and Def-2 [kappa (SE)=0.59 (0.05)]. A substantial agreement was found between Def-1 and the self-reported PEx [kappa (SE)=0.61 (0.05)]. Between Def-2 and the self-reported PEx the agreement was moderate [kappa (SE)=0.51 (0.05)] (Table 3).

**Table 3:** Cohen’s kappa statistic test for the agreement between different exacerbation definitions within a year in n=248 participants with genetically confirmed PCD; Def-1: Definition 1, Def-2: Definition 2

|  | Kappa | Std. error | Agreement |
| --- | --- | --- | --- |
| Def-1 vs Def-2 | 0.59 | 0.05 | moderate |
| Self-reported vs Def-1 | 0.61 | 0.05 | substantial |
| Self-reported vs Def-2 | 0.51 | 0.05 | moderate |

### Risk factors for PEx in PCD

For participants included in the annual analysis (n=248), a mixed effects model, with a random effect for the centre, showed that female sex increased the risk of PEx independent of the definition used (for self-reported PEx: p= 0.039; for Def-1: p=0.012) and Def-2 (p=0.047) (Table 4). Age was positively associated with higher PEx frequency with Def-1 (p=0.034), but not with Def-2 (p=0.863) and self-reported PEx (p=0.925). Furthermore, we found a positive, but not statistically significant association between isolation of *Pseudomonas aeruginosa* at least once during the study period and PEx frequency with Def-2 (p=0.057), but not with the other two definitions. FEV_1_ z-score was not associated with PEx frequency, independent of the PEx definition.

**Table 4:**
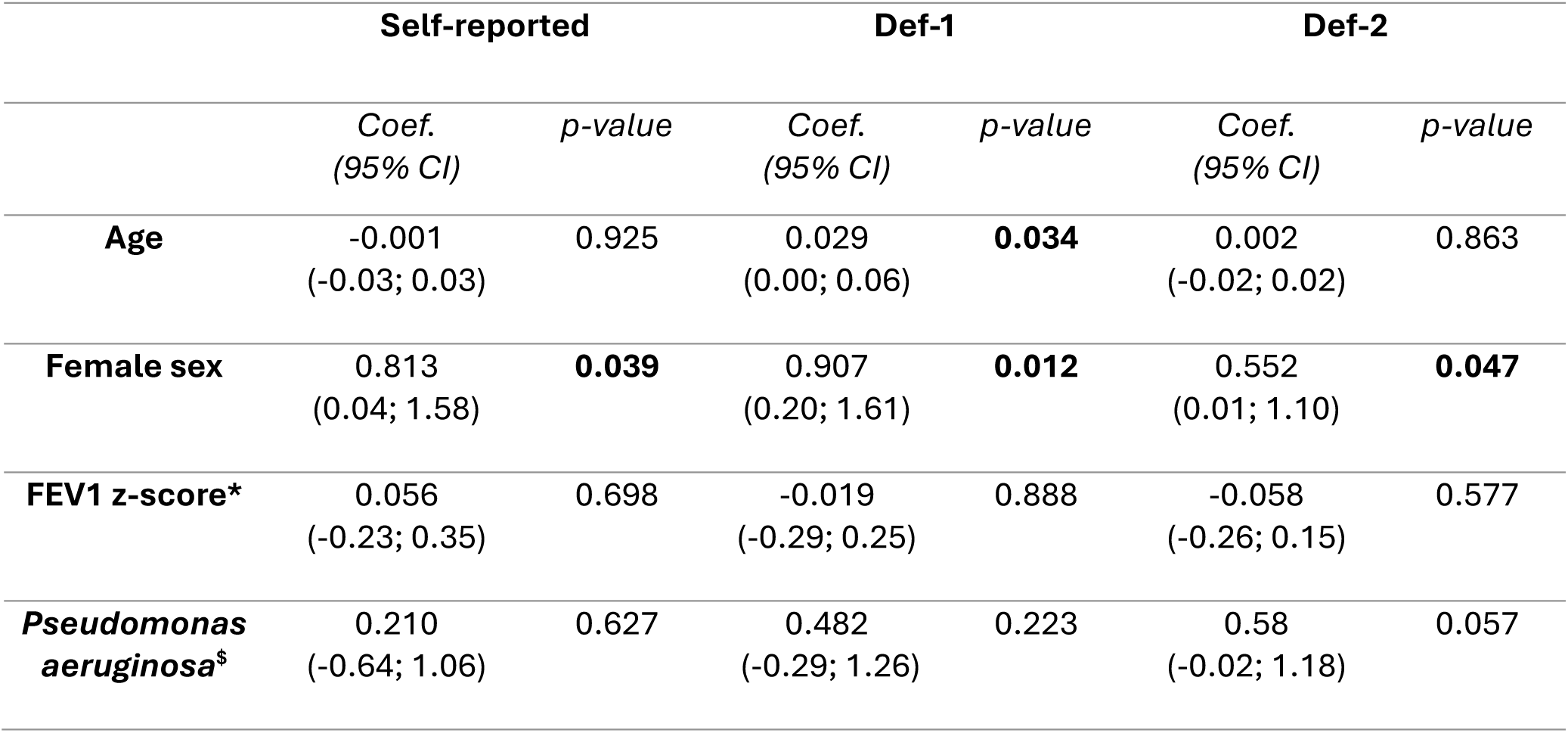
The role of age, sex, lung function and Pseudomonas aeruginosa in PEx frequency by each definition, in n=248 participants with genetically confirmed PCD and complete annual records. Mixed effects model with random effect for the center for the three exacerbation definitions; Def-1: Definition 1, Def-2: Definition 2; * Median FEV_1_ (forced expired volume at 1 sec) during the study period; ^$^ At least once positive culture during the study period

|  | Self-reported |  | Def-1 |  | Def-2 |  |
| --- | --- | --- | --- | --- | --- | --- |
|  | Coef.<br>(95% CI) | p-value | Coef.<br>(95% CI) | p-value | Coef.<br>(95% CI) | p-value |
| **Age** | -0.001<br>(-0.03; 0.03) | 0.925 | 0.029<br>(0.00; 0.06) | **0.034** | 0.002<br>(-0.02; 0.02) | 0.863 |
| **Female sex** | 0.813<br>(0.04; 1.58) | **0.039** | 0.907<br>(0.20; 1.61) | **0.012** | 0.552<br>(0.01; 1.10) | **0.047** |
| **FEV1 z-score\*** | 0.056<br>(-0.23; 0.35) | 0.698 | -0.019<br>(-0.29; 0.25) | 0.888 | -0.058<br>(-0.26; 0.15) | 0.577 |
| ***Pseudomonas aeruginosa*<sup>§</sup>** | 0.210<br>(-0.64; 1.06) | 0.627 | 0.482<br>(-0.29; 1.26) | 0.223 | 0.58<br>(-0.02; 1.18) | 0.057 |

### Impact of age on PEx frequency

Considering that airway infections are more common in children compared to adults, we performed a subgroup analysis by grouping participants by age into children (0-17.9 years) and adults (≥18 years). Interestingly, we found no significant difference between Def-1 and Def-2 (p=0.65) in children. In adults, PEx frequency as defined by Def-2 was statistically lower compared to self-reported (p<0.001) and Def-1 (p=0.004) (Suppl. Figure S2 and suppl. Table S1).

For children ≤12 years it was the caregiver that provided the relevant information during the telephone interview, while all participants >12 years responded by themselves. We aimed to study whether this difference had any systematic effect on the results. The annual PEx frequency per person was significantly higher in children ≤12 years compared to children 12.1-17.9 years for self-reported PEx (p=0.003), for Def-1 (p=0.028) and for Def-2 (p=0.029) (Supplementary Table S2).

### Genetic diagnosis and PEx frequency

Participants were further separated according to their genetic diagnosis to those predicted to present Class I(n=175) or Class II(n=72) defects in cilia ultrastructure (Supplementary Table S3). No difference was found in PEx frequency between the two groups, independent of the definition used. In both groups, self-reported PEx were significantly higher compared to those assessed with Def-1 (p<0.0001) and Def-2 (p<0.001). Only among patients predicted to present Class I defects we found a significant difference between Def-1 and Def-2 (p=0.041).

### Geographical impact on PEx frequency

We attempted a geographical subgrouping between centers located in the central/northern Europe (Denmark and Germany) and southern Europe (Spain, Italy, Greece, Cyprus, Turkey). Participants from central/northern Europe (n=72) had significantly higher annual PEx frequency in all three definitions, compared to participants from southern Europe (n=176), although there were no age differences between the two groups (Supplementary Table S4).

### Seasonal distribution of PEx

Analysis of PEx by season in all study participants (n=271) showed that the highest frequency of exacerbations occurred in autumn when using self-reports (OR: 1.47, 95% CI: 1.16-1.87, p=0.001), with Def-1 (OR: 1.40, 95% CI: 1.08-1.83, p=0.012), and with Def-2 (OR: 1.44, 95% CI: 1.10-1.88, p=0.007), while the lowest frequency was recorded in summer (Figure 2). The same seasonal pattern was observed when we analysed separately i) children and adults, and ii) participants from southern and central/ northern Europe. Examination of the monthly distribution of PEx revealed that October and November were the months with more frequent PEx (35.24% and 35.47% of patients with self-reports, 24.07% and 25.37% with Def-1, 20.34% and 21.67% with Def-2, respectively). A sensitivity analysis that included only n=248 participants with complete annual records showed the same distribution pattern.

**Figure 2:**
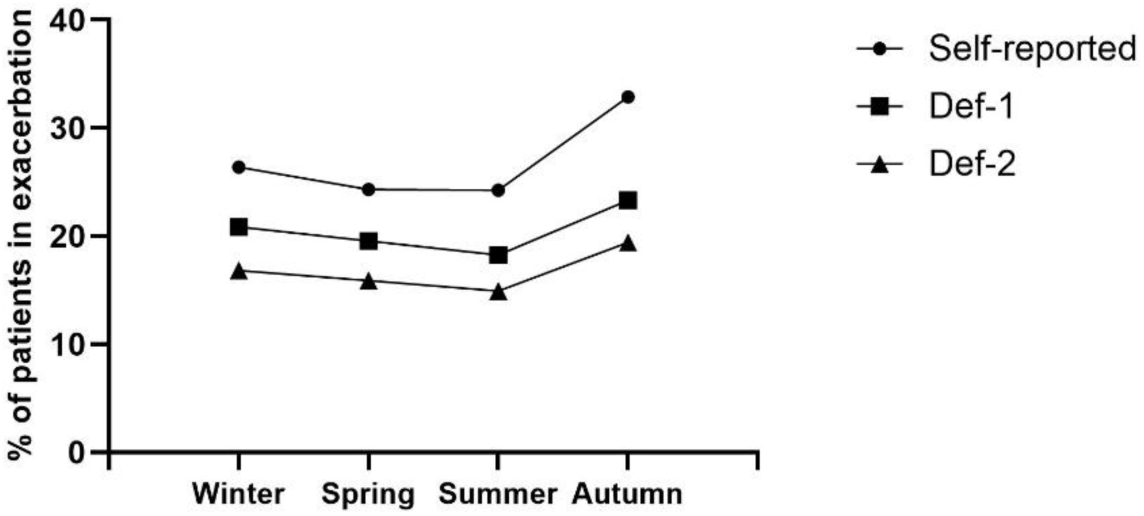
Seasonal distribution of pulmonary exacerbation frequency in n=271 patients with genetically confirmed PCD. Self-reported: self-reported exacerbations, Def-1: Definition 1, Def-2: Definition 2

## DISCUSSION

This is a prospective multicentre study specifically designed to assess the annual frequency of PEx in an existing well-structured cohort of patients with genetically confirmed PCD from the International PCD Registry, within the ERN LUNG PCD Core. For the first time two most common definitions of PEx in PCD found in the literature, and the patients’ personal perception of experiencing PEx were used towards this goal.

The annual PEx frequency was highly variable within our cohort. Within one year, over 30% of study participants had at least three PEx, whereas approximately 20% had no exacerbation. This finding correlates well with previous studies showing high heterogeneity of the disease manifestations and symptoms severity [1] [2] [3] [4]. Because of the high variability, we chose to report results as median values rather than mean values or incidence rates, although the incidence rate of annual exacerbations is by definition higher.

The present study found a moderate agreement in PEx frequency between the two existing PEx definitions. This could be partially attributed to the different approach each definition uses to determine PEx. For Def-1, PEx is recorded when three out of seven clinical items are positive. Of these, only two (start/change of antibiotic treatment and fever>38 ^0^C) are clearly objective, while the others (increased cough, increased sputum, dyspnoea, fatigue, hemoptysis) are qualitatively and subjectively assessed. This may introduce a degree of bias in the definition, which to some extent depends on the patient’s (or caregiver’s) perception. On the other hand, Def-2 uses exclusively antibiotic treatment to determine a PEx, which is more pragmatic and easier to assess. However, it has the risk of relying on patients’ attitude towards therapies and “medical advice seeking behavior”. It also depends on the prescribing practices of healthcare providers. Interestingly, we found no difference between the two definitions in the pediatric population of the study. In contrast, adult patients had a higher rate of PEx by Def-1 compared to Def-2. This suggests that in children with PCD, both parents and healthcare professionals are more eager to administer antibiotics for exacerbations, compared to older patients. Taken together, our study does not demonstrate superiority of either clinical PEx definition, although their use is not interchangeable.

Previous single-center studies with fewer patients report either similar or lower PEx annual frequency, primarily assessed by Def-1 [18] [19]. However, the study design was different and the diagnosis of PCD was not always confirmed, thus a direct comparison cannot be guaranteed. A recent international study, which used data self-provided by patients with PCD on a patient-initiated online platform, indicated a slightly higher incidence rate of PEx by Def-1 [34]. As the participation in this study has been determined by the patients’ initiative, it might represent a subgroup of patients with particular concern about their health. Also, diagnostic details were not available for the whole cohort, which increases the risk to include patients not fulfilling the diagnostic criteria for PCD.

The present study captured an important parameter that is often neglected in clinical research, the patient-reported exacerbation rate, highlighting the discordance between the patient’s perception of PEx and existing clinical definitions. Although clinical metrics and cutoffs are necessary to define a PEx, there is a possibility that they cannot capture entirely the burden of this condition. The impact of worsening symptoms on the quality of everyday life is often subjective and may exceed clinical definitions. Notably, Def-1, which uses several qualitative measures, had slightly better agreement with self-reports compared to Def-2, suggesting that it might be closer to patients’ perception on exacerbations. In COPD, the frequency of exacerbations was found to be higher in studies that record the patients’ perception on PEx compared to those using clinical PEx definitions [27] [28]. These data further support the need to include patient-reported outcomes in future clinical trials.

Our analysis revealed several risk factors for PEx. Independent of the definition used, the annual PEx frequency was higher in females than in males. This finding has been also described in the study of Schreck et al [34]. Interestingly, female sex has been reported as a risk-factor for increased exacerbations in other chronic lung diseases, such as cystic fibrosis [35] [36] and COPD [37]. Further studies are needed to explore possible mechanisms behind this discrepancy in PCD. An age-dependent increase in PEx was observed in Def-1. This could be expected given the age-related progression of chronic lung disease in PCD that is often complicated by the development of bronchiectasis. However, this was not observed with Def-2, probably because adult patients do not always use antibiotics during an exacerbation. Subgroup analysis by age showed that children up to the age of 12 had more frequent PEx than teenagers (12-18 years). We hypothesize that the increased prevalence of viral infections in younger children may play a role in this difference [38], but an overestimation of symptoms by the parents at this age cannot be excluded. However, as this finding applies also to Def-2, which corresponds to an objective question (the use of antibiotics), it cannot be solely attributed to differences between patients and caregivers in providing the relevant information.

Isolation of *Pseudomonas aeruginosa* has been identified as a risk factor for PEx in PCD [15] [34] and other chronic lung diseases [39]. In our study there was a positive, but not significant, association between PEx, as defined by Def-2, and at least once positive airway culture for *Pseudomonas aeruginosa*. More targeted studies are needed to investigate in depth the role of microbial and viral airway abundance, as well as environmental exposures as risk factors for PEx. FEV1 was not associated with PEx frequency, but this might be attributed to the high percentage of the study participants (66.5%) having FEV_1_ levels above the lower limit of normal (Table 2) at the time of the study.

Data recordings throughout the year revealed seasonal differences in PEx. Autumn appears to be the season that burdens the most patients with PCD in terms of PEx, regardless of the definition used. This pattern has been observed in both patients from central/northern and southern Europe, although the first group had a higher annual PEx frequency. There is a rise in PEx during October and November, which may be due to the high prevalence of viral respiratory infections during this time of the year, considering that all participating centres are located in the northern hemisphere. In addition, possible changes in exposure to aeroallergens and air pollutants as well as climatic changes occurring during this period of the year could also increase the susceptibility to exacerbations of patients with obstructive lung disease [40]. A similar seasonal pattern has been described in asthmatics [41]. This is an emerging area of research in PCD. Future clinical studies should address the above issues in more detail [42], and evaluate possible mitigation strategies to reduce the burden of exacerbations during high-risk periods.

Our study has several strengths. A well-described patient cohort meeting robust diagnostic criteria [23] based on genetics for PCD from multiple centres and countries was included in the analysis, which supports the validity and transparency of the reported outcomes. Data were systematically collected monthly for at least 12 months by healthcare professionals using standardised interviews. Complete annual data were included to calculate the annual exacerbation frequency. The study was inclusive by design, regardless of the participants’ clinical status and frequency of clinical visits. Telephone contacts encouraged patients who did not consistently attend outpatient clinics to participate in the study, who would otherwise be missed in a clinical setting. The COVID-19 quarantine period (during 2020) has been associated with lower exacerbation rates in PCD [19] and was excluded from this analysis, although self-protective behaviours by some patients might have continued during 2021 and 2022 leading to an underestimation of the PEx frequency. Furthermore, we were able to evaluate the two existing PEx definitions simultaneously in each participant and record the monthly/seasonal distribution.

On the other hand, the results should be interpreted in consideration of several limitations. Data were collected from patient/caregiver interviews and do not include physician confirmation of exacerbations. They also lack information on the severity of exacerbations. In addition, neither the questionnaire used in this study, nor the Def-1 have been so far validated. The clinical data were collected in parallel but not always synchronously to the monthly questionnaire, so the microbiological profile of each patient could not be directly related to the reported exacerbations. In addition, due to the current format of the International PCD Registry, there is no information on chronic microbial colonisation or an exhaustive input of microbiological findings. Last, our cohort includes only a small number of patients over 50 years of age (7.7%), so our results may not be representative of this age group.

In conclusion, this is the first multicentre study that prospectively evaluated the frequency of PEx in a large international cohort of patients with a genetically confirmed diagnosis of PCD. We demonstrated the high heterogeneity in the incidence of PEx and highlighted the importance of the exacerbation definition in capturing the burden of the disease, as well as the influence of biological sex, age, and season on the exacerbation rate. Clinical researchers and the pharmaceutical industry should consider both the study objectives and the population characteristics before selecting the most appropriate PEx definition for future clinical trials [43].

Part of these results has been presented as a poster at the European Respiratory Society Conference 2024 in Vienna, Austria (Anagnostopoulou P, *et al*. European Respiratory Journal 2024 64(suppl 68): PA2382; DOI: https://doi.org/10.1183/13993003.congress-2024.PA2382)

## ACKNOWLEDGEMENTS

The authors would like to thank the participants with PCD and their families for taking part in the study. They also acknowledge the work of S. Philippou, S. Hadjievagorou, A. Anagnostopoulou, S. Louka, A.M. Sahtouri, A. Avraam, C. Antoniou, S. Cheilidis, K. Sykallos who performed the data-cleaning. This work is generated within the European Reference Network for Rare Respiratory Diseases (ERN-LUNG). Several authors are part of the Beat-PCD Clinical Research Collaboration.

## Author contributions

PA: project management, study design, study monitoring, data curation, data analysis, manuscript preparation, PK: study design, data curation, data analysis, manuscript preparation, DAT: data acquisition, SA: data acquisition, SBQ: data acquisition, MB: data acquisition, AC: data acquisition, HNC: data acquisition, RC: data acquisition, NE: data acquisition, EE: data acquisition, YG: data acquisition, SG: data acquisition, EH: local-PI, data acquisition, SH: data acquisition, data curation, BK: local-PI, data acquisition, DM: data acquisition, MAM: local-PI, study design, manuscript preparation, JKM: study design, manuscript preparation, NM: data analysis, AM-G: local-PI, data acquisition, manuscript preparation, UO: local-PI, data acquisition, MP: local-PI, data acquisition, JR: study design, manuscript preparation, JRö: data acquisition, study design, manuscript preparation, SR-A: data acquisition, FS: local-PI, data acquisition, study design, manuscript preparation, IT: data acquisition, NU: local-PI, data acquisition, NZ: data acquisition, KGN: local-PI, data acquisition, study design, manuscript preparation, HO: local-PI, study design, manuscript preparation, PY: study design, data analysis, manuscript preparation. All authors read and approved the submitted manuscript.

## Data availability statement

All data that support the findings of this study are available from the International PCD Registry, but restrictions apply to the availability of these data, which were used under licence for the current study and so are not publicly available. The data are, however, available upon request and with the permission of the data contributors.

