## Supplementary data for "Pulmonary exacerbations in patients with genetically confirmed PCD: A prospective observational multicentre study"

#### Methods

##### Participating centers

The participating centers in the study are listed below:

- Medical School, University of Cyprus & Paediatric Pulmonology Unit, Archbishop Makarios III Hospital, Cyprus
- Danish PCD Centre Copenhagen, Paediatric Pulmonary Service, Copenhagen University Hospital, Denmark
- Department of General Pediatrics, University Children's Hospital Muenster, Muenster, Germany
- Clinic for Pediatrics, Pneumology, Immunology and Intensive Care Medicine, Charite Universitätsmedizin Berlin, Berlin Germany
- Department of Pediatrics, Vall d'Hebron Hospital Universitari, Vall d'Hebron Barcelona Hospital Campus, Universitat Autònoma de Barcelona, Barcelona, Spain
- Pediatric Pulmonology and Respiratory Intermediate Care Unit, Sleep and Long-Term Ventilation Unit, Academic Department of Pediatrics, Bambino Gesù Children's Hospital, Rome, Italy
- Department of Paediatrics, University Hospital of Pisa, Pisa, Italy
- Department of Translational Medical Sciences, Pediatric Pulmonology, Federico II University, Naples, Italy
- Hacettepe University, Department of Pediatric Pulmonology Ankara, Turkey
- Pediatric Pulmonology Unit, 3rd Pediatric Clinic, Hippokrateio General Hospital, Thessaloniki, Greece
- Department of Pediatric Pulmonology, Marmara University, School of Medicine, Istanbul, Turkey

**Genetic diagnosis**

Participants' genetic data were retrieved from the ERN-LUNG International PCD Registry. Genetic variants were evaluated according to American College of Medical Genetics and Genomics (ACMG)/Association for Molecular Pathology (AMP) guidelines [S1]. Only variants classified as pathogenic (ACMG/AMG class 5) or likely pathogenic (ACMG/AMG class 4) were considered as disease-causing and included for further analysis [S2].

**Data collection**

Data collection was initially planned for the years 2020 and 2021 but due to the COVID-19 pandemic, was extended up to the end of 2022 and analysed the last 12 months of each patient's participation in the study.

- Monthly structured telephone interviews using a standardised questionnaire

The questionnaire was developed in English for the needs of the study, taking into consideration the published PEx definitions [S3] [S4] [S5] [S6], and subsequently translated into the official language of each centre using a translation-back translation protocol [S7]. A trained healthcare professional conducted monthly a telephone interview with the patient or the caregiver for children ≤12 years and completed the questionnaire. The telephone calls were organised by each centre, and the completed questionnaires were uploaded to the International PCD Registry platform [S8]. To minimise the inter-rater bias the colleagues that conducted the telephone interviews were trained in group meetings by the study coordinator (PA) in several time-points before and during the study. Further, the centers were advised to appoint one person that conducts all the telephone interviews of the same patient during the study.

- Clinical data

We used data (anthropometrics, sputum/pharyngeal swab microbiology, spirometry) collected in parallel routine and emergency onsite clinical visits in each centre during the study period (including 3 months before the beginning of the study and 3 months after the end of the study) that were uploaded to the International PCD Registry. For patients with multiple spirometric measurements during the study period, we used the median forced expiratory volume in 1 sec ($FEV_1$) z-score. According to the data collection requirements of the International PCD Registry, the available microbiology results correspond only to the above-mentioned period, therefore at least one positive culture per participant is reported, whereas no information on chronic bacterial colonisation was available.

Data extraction from the International PCD registry platform was performed at the end of the study.

| | **Self-reported** | **Def-1** | **Def-2** |
|---|---|---|---|
| **Children (n=127)** | 3 (1-5) | 1 (0-3) | 2 (0-3) |
| **Adults (n=121)** | 2 (1-4) | 2 (0-4) | 1 (0-3) |
| **p-value*** | 0.082 | 0.653 | 0.687 |

Supplementary Table S1: Annual pulmonary exacerbation frequency in children (0-17.9 years) and adults (≥18 years) with genetically confirmed PCD, as defined by Self-reports, Definition-1 (Def-1) and Definition-2 (Def-2). Results presented as median (interquartile range); * Mann-Whitney test.

| | **Self-reported** | **Def-1** | **Def-2** |
|---|---|---|---|
| **0-12 years (n=67)** | 4 (1-6) | 2 (1-4) | 2 (1-4) |
| **12.1 -17.9 years (n=60)** | 2 (1-4) | 1 (0-2.75) | 1 (0-2) |
| **p-value*** | **0.003** | **0.028** | **0.029** |

Supplementary Table S2: Annual pulmonary exacerbation frequency in children <6 years) and school-aged children (6-<18 years) with genetically confirmed PCD, as defined by Self-reports, Definition-1 (Def-1) and Definition-2 (Def-2). Results presented as median (interquartile range); * Mann-Whitney test.

| | Self-reported | Def-1 | Def-2 |
|---|---|---|---|
| **Class I# (n=175)** | 2 (1-5) | 2 (1-4) | 2 (1-3) |
| **Class II$ (n=73)** | 3 (1-5.5) | 1 (0-4) | 1 (0-4) |
| **p-value*** | 0.576 | 0.749 | 0.789 |

Supplementary Table S3: Annual pulmonary exacerbation frequency in participants who, according to their genetic diagnosis, are predicted to present Class I or Class II defects in cilia ultrastructure, see ref. [25], as defined by Self-reports, Definition-1 (Def-1) and Definitions-2 (Def-2). Results presented as median (interquartile range);

### bi-allelic variants in: *DNAH5, DNHA9, DNAI1, DNAI2, ARMC4/ODAD2, CCDC114/ODAD, CCDC151, TTC25/ODAD4, CCDC103, DNAAF7/ZMYND10, DNAAF4/DYX1C1, DNAAF3/C19ORF51, CFAP300/C11ORF70, DNAAF11/LRRC6, DNAAF1/LRRC50, SPAG1, CCDC40, CCDC39;* hemizygous variants (males) in: *DNAAF6/PIH1D3* (X-linked recessive inheritance)

$ bi-allelic variants in: *HYDIN, SPEF2, DRC1/CCDC164, RSPH4A, RSPH1, RSPH9, DNAH11, CCNO, DNAJB13*; heterozygotes in *FOXJ1* (autosomal dominant inheritance), hemizygous variants (males) in *RPGR, OFD1* (X-linked recessive inheritance)

* Mann-Whitney test

| | Self-reported | Def-1 | Def-2 |
|---|---|---|---|
| **Central/Northern Europe (n=72)** | 4 (1-8) | 3 (1-6) | 2 (1-4) |
| **Southern Europe (n=176)** | 2 (1-4) | 1 (0-3) | 1 (0-2) |
| **p-value*** | **<0.001** | **<0.001** | **<0.001** |

Supplementary Table S4: Pulmonary exacerbation frequency per year in participants form central/northern Europe (Germany, Denmark) and participants from Southern Europe (Spain, Italy, Greece, Cyprus, Turkey) with genetically confirmed PCD, as defined by Self-reports, Definition-1 (Def-1) and Definition-2 (Def-2). Results presented as median (interquartile range); * Mann-Whitney test.

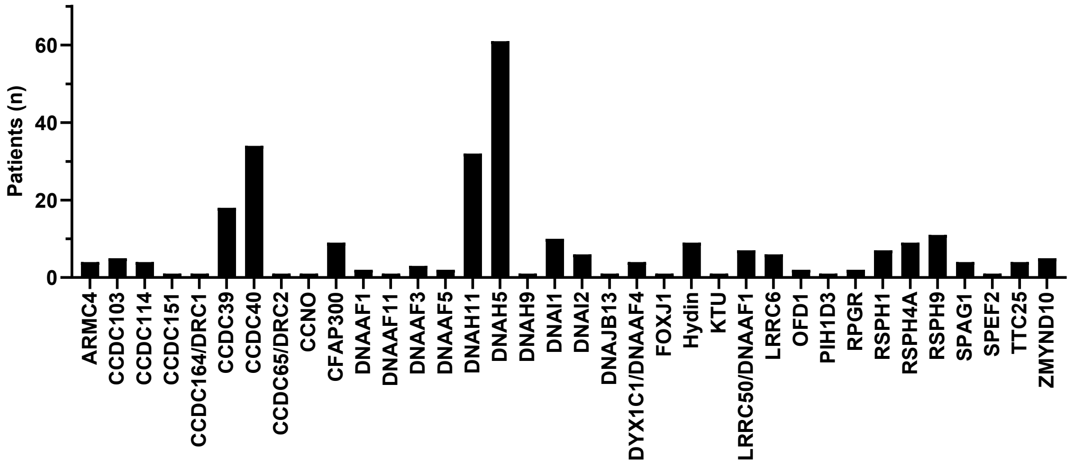


Supplementary Figure S1: Distribution of study participants with pathogenic variants in PCD-causing genes (n=248)

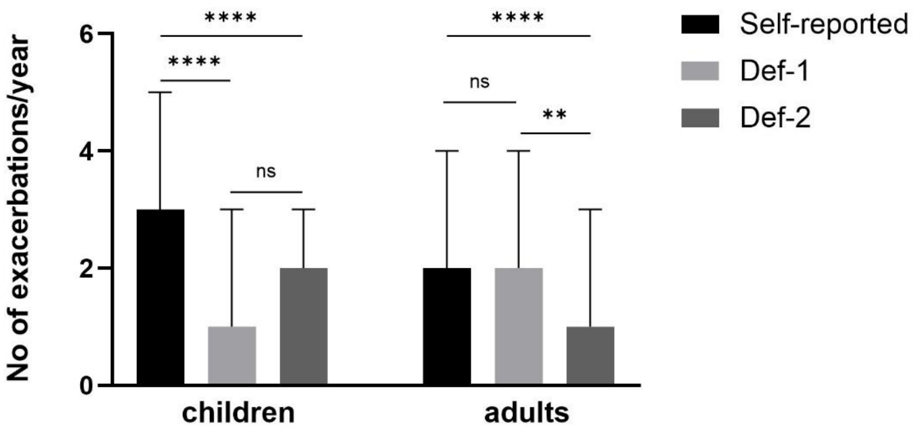


Supplementary Figure S2: Annual frequency of pulmonary exacerbations as recorded by self-reports (Self-reported), Definition 1 (Def-1) and Definition 2 (Def-2) in children (0-17.9 years) and adults (≥18 years) with genetically confirmed PCD. Results are shown as median (interquartile range). For the comparison of different definitions within the same group: Wilcoxon-matched pairs test, **** p<0.0001, ** p<0.01, ns: non-significant.
